# Development and Validation of the VIOSync Sepsis Prediction Index: A Novel Machine Learning Model for Sepsis Prediction in ICU Patients

**DOI:** 10.1101/2024.02.22.24303211

**Authors:** Sotirios G. Liliopoulos, Alexander Dejaco, Vasileios S. Dimakopoulos, Ioannis A. Gkouzionis

## Abstract

**Background:** Sepsis is the third leading cause of death worldwide and the main cause of in-hospital mortality. Despite decades of research, sepsis remains a major challenge faced by patients, clinicians, and medical systems worldwide. Early identification and prediction of patients at risk of sepsis and adverse outcomes associated with sepsis are critical. In this work, we aimed to develop an artificial intelligence algorithm that can predict sepsis early.

**Materials and Methods:** We developed a predictive model for sepsis using data from the Physionet Cardiology Challenge 2019 ICU database. Our cohort consisted of adult patients who were admitted to the ICU. Sepsis diagnoses were determined using the Sepsis-3 criteria. The model, built with the XGBoost algorithm, was designed to anticipate sepsis prior to the appearance of clinical symptoms. An internal validation was conducted using a hold-off test dataset to evaluate the AI model’s predictive performance.

**Results:** We have developed the VIOSync Sepsis Prediction Index (SPI), an AI-based predictive model designed to forecast sepsis up to six hours before its clinical onset, as defined by Sepsis-3 criteria. The AI model, trained on a dataset comprising approximately 40,000 adult patients, integrates variables such as vital signs, laboratory data, and demographic information. The model demonstrated a high prediction accuracy rate of 97%, with a sensitivity of 87% and a specificity of 98% in predicting sepsis up to 6 hours before the onset. When compared to the established qSOFA score, which has a specificity of 89% for sepsis prediction, our VIOSync SPI algorithm significantly enhances predictive reliability, potentially reducing false positive rates by a factor of 5.5.

**Conclusions:** The VIOSync SPI demonstrated superior prediction performance over current sepsis early warning scores and predictive algorithms for sepsis onset. To validate the generalizability of our method across populations and treatment protocols, external validation studies are essential.

## INTRODUCTION

Sepsis is identified as a critical condition characterized by life-threatening acute organ dysfunction caused by a dysregulated host response to infection (Singer *et al*., 2016). Recognizing the gravity of sepsis, in 2017, global health organizations including the World Health Assembly and the World Healthcare Organization prioritized its detection, prevention, and treatment globally (Reinhart *et al*., 2017; Paoli *et al*., 2018). It is estimated that sepsis affects 4-6% of adult hospital admissions (Rhee *et al*., 2017; Giamarellos-Bourboulis *et al*., 2023; Mellhammar *et al*., 2023) and is found in about one-third of patients in intensive care units (ICU) (Sakr *et al*., 2018). In 2017 alone, nearly 49 million people globally were affected by sepsis, with 11 million succumbing to the condition, indicating a mortality rate of about 20% (Rudd *et al*., 2020). Particularly, in the United States, there are approximately 1.7 million cases for sepsis per year, a trend that has been increasing annually. This condition results in nearly 250,000 deaths annually in the U.S. alone, making sepsis the primary cause of mortality in non-cardiac ICUs (Vincent *et al*., 2009; Rhee *et al*., 2017). Despite the steady admission rate of sepsis patients to ICUs across European hospitals from 2002 to 2012, the severity of the disease increased significantly (Vincent *et al*., 2018). Mortality rates vary widely but are reported to be at least 10%, jumping to 40% in cases involving septic shock (Vincent *et al*., 2018), and exceeding 30% when sepsis is left untreated (Liu *et al*., 2014; Rhee *et al*., 2017). Additionally, the financial burden of sepsis treatment is substantial. In the U.S., hospital expenses for sepsis management were the highest among all diseases, exceeding USD 20 billion in 2011, reaching over USD 23 billion in 2013, and consistently costing more than USD 24 billion annually, which represents 13% of total U.S. healthcare expenditures (Arefian *et al*., 2017; Reinhart *et al*., 2017; Paoli *et al*., 2018; Buchman *et al*., 2020).

Prompt and effective intervention for sepsis is critical, particularly in ICUs, where the most critically ill patients are treated. The urgency in treating sepsis cannot be overstated, as mortality rates increase by approximately 4-8% with each hour treatment is delayed (Churpek *et al*., 2016). Studies have shown that early identification of sepsis can minimize treatment delays, enhance the delivery of suitable interventions, and ultimately reduce mortality (Kumar *et al*., 2006; Mok *et al*., 2014; Husabø *et al*., 2020). Despite the high risk of mortality associated with sepsis, there is general consensus in medical guidelines (Rhodes *et al*., 2017) that prompt action involving antibiotics, fluid resuscitation, source control, and support of vital organ function lead to dramatically improved patient outcomes. The challenge in early sepsis detection lies in its heterogeneous syndromic nature, which can evolve based on diverse pathophysiological factors, the complexity of each clinical case, and the clinical phenotypes. This challenge is compounded by the absence of reliable blood- or plasma-based biomarkers for early detection of sepsis. Although hundreds of potential biomarkers have been evaluated for their prognostic value in sepsis (Pierrakos and Vincent, 2010; Cho and Choi, 2014; Pierrakos *et al*., 2020), their lack of sufficient specificity or sensitivity prevents their routinely use in clinical practice (Pierrakos and Vincent, 2010). There is thus a significant unmet need for new tools to support clinicians swiftly identifying hospitalized patients at risk of developing sepsis.

Currently, sepsis diagnosis involves a combination of clinical assessments by healthcare professionals and data from monitoring devices and screening laboratory tests. This approach is both time-intensive and subjective, relying heavily on the expertise and judgment of the healthcare professional. Timely intervention is critical for patients with sepsis, yet with the manual routines used at present, there is a risk of delayed diagnosis of sepsis and initiation of treatment. Enhancing the timely prediction and detection of patients at risk of developing sepsis is crucial for mitigating its detrimental effects. Given the complexity of sepsis as a clinical syndrome, characterized by a broad spectrum of clinical and biological indicators, relying on a single clinical marker may not accurately represent the disease’s state (Hernandez, Bellomo and Bakker, 2019).

To capture the critical window for controlling sepsis progression, clinical practices often implement rule-based scoring systems, such as the systemic inflammatory response syndrome (SIRS) criteria (van Wyk *et al*., 2019), sequential organ failure assessment (SOFA) scores (Vincent *et al*., 1996), and modified early warning score (MEWS), to alert the possible occurrence of sepsis. The timely application of these scoring methods facilitates early detection and allows for the initiation of preemptive treatment measures or alert programs with a high degree of sensitivity. Nevertheless, while these systems are effective in identifying potential sepsis cases, they often lack specificity, leading to false alarms and thus alert fatigue.

Given that intensivists are overwhelmed with the ever-increasing volume of data collected at the bedside, the interest in machine learning prediction algorithms has surged within both research and clinical practice. This growing attention is attributed to the algorithms’ potential to enhance early detection, ensure better compliance with treatment protocols, and reduce the time to antibiotic administration. Such improvements have been demonstrated to significantly improve patient outcomes (Kumar *et al*., 2006; Mok *et al*., 2014; Seymour *et al*., 2017; Husabø *et al*., 2020). To date, and to the best of our knowledge, three ICU algorithms are available for clinical use (Henry *et al*., 2015; Calvert *et al*., 2016; Persson *et al*., 2021). Persson et al. leveraged the MIMIC-III database to devise the NAVOY algorithm, which forecasts sepsis occurrence within ICU settings up to 3 hours prior to its onset (Persson *et al*., 2021). Calvert et al. utilized the MIMIC-II database to create the InSight algorithm, also predicting sepsis onset with a 3-hour lead time (Calvert *et al*., 2016). In a similar vein, Henry et al. analyzed physiological and laboratory data from ICU patients, resulting in the development of TREWScore, a predictive tool capable of anticipating septic shock up to 28 hours in advance (Henry *et al*., 2015). Nemati et al. used electronic medical record data combined with high-resolution time series of heart rate and blood pressure to dynamically predict sepsis, with an area under the receiver operating characteristic (AUROC) of 0.83–0.85 (Nemati *et al*., 2018). However, a limitation among these studies is the lack of information on potential confounding factors or variables not included in the predictive models that could influence the accuracy of sepsis prediction within ICU settings.

The purpose of this study is to harness the potential of machine learning technologies to address a critical challenge in ICUs worldwide: the early prediction of sepsis. By leveraging clinical data routinely collected in electronic health records (EHRs), this study develops a sophisticated machine learning algorithm capable of predicting the onset of sepsis within a crucial six-hour window. The adoption of machine learning in healthcare offers a promising avenue for enhancing predictive analytics, surpassing traditional statistical methods in accuracy, speed, and efficiency. The successful development of such an algorithm has the potential to significantly impact patient care in ICUs by enabling timely interventions, thereby reducing mortality rates, improving overall patient outcomes, and optimizing the allocation of healthcare resources.

## MATERIALS AND METHODS

In addressing the critical challenge of early sepsis detection in ICU patients, our study leverage advanced machine learning techniques to develop a predictive model capable of accurately predicting sepsis before onset. The careful selection and fine-tuning of these techniques are pivotal in ensuring the model’s practical applicability and reliability in clinical environments. For this reason, we have developed an exhaustive methodology that harmonizes clinical dat examination with advanced algorithmic approaches (**Figure 1**). All computational analyses and model development were conducted using Python.

**Figure 1.**
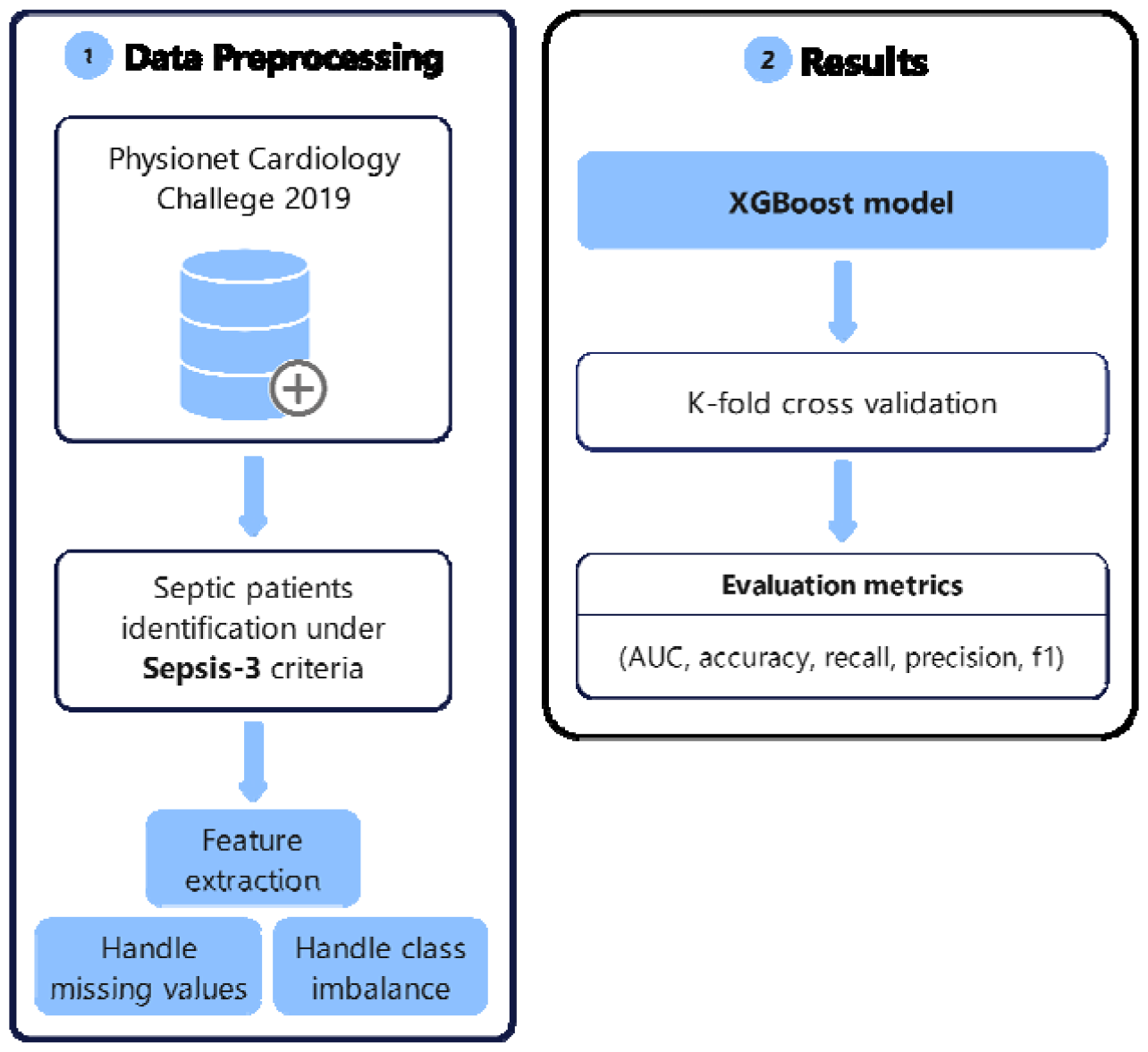
Flowchart of the proposed sepsis prediction workflow from preprocessing datasets to evaluating sepsis predictions using XGBoost.

### Dataset and Study Population

We used the PhysioNet/Computing in Cardiology Challenge 2019 dataset (Goldberger *et al*., 2000; Reyna *et al*., 2020). The data were obtained from three geographically distinct U.S. hospital systems with three different Electronic Medical Record (EMR) systems: Beth Israel Deaconess Medical Center, Emory University Hospital, and a third, unidentified hospital system. These data were collected over the past decade with approval from the appropriate institutional review boards. Data and sepsis labels from a total of 40,336 patients were used. The data consisted of a combination of hourly vital sign observations, laboratory values, and static patient descriptions. The data contained 40 clinical variables: eight vital sign variables, such as heart rate, respiration rate, blood pressure and blood oxygen saturation, 26 laboratory variables (e.g., lactate, bilirubin, hemoglobin, etc.), and six demographic variables such as age, gender, and ICU length-of-stay (hours since ICU admit). Altogether, these data included over 2.5 million hourly time windows and 15 million data points.

### Data Labeling

The data was labeled in accordance with Sepsis-3 criteria (Singer *et al*., 2016). Three time points were specified for each septic patient in order to define the onset time of sepsis. These include tsuspicion, marking the initial suspicion of infection based on the administration of intravenous (IV) antibiotics and blood culture timings; tSOFA, indicating the occurrence of organ failure as evidenced by a two-point increase in the SOFA score within 24 hours; and tsepsis, defined as the onset of sepsis, determined by the earliest tsuspicion or tSOFA, as long as tSOFA occurs no more than 24 hours before or within 12 hours after tsuspicion. These criteria ensured that IV antibiotics are administered for a minimum of 72 consecutive hours, with a mandatory temporal relationship between the administration of IV antibiotics and the acquisition of blood cultures to accurately reflect clinical practice (Singer *et al*., 2016). This approach characterized septic patients as those with a finite tsepsis, whereas non-septic patients were characterized by an infinite tsepsis value. Septic patients were assigned a label=1, while non-septic patients received a label=0, providing a binary classification framework for our analysis. To enhance the predictive model’s utility in clinical settings by enabling early intervention, labels for septic patients were shifted ahead by six hours, indicating the goal to predict sepsis onset six hours before it clinically manifests. This temporal adjustment allows the model to identify potential sepsis cases with a lead time, offering a crucial window for preemptive medical intervention. This approach provides a clear, standardized method for labeling and analyzing patient data in the predictive modeling of sepsis.

### Handling Missing Values

A critical aspect of preparing the dataset for the early prediction of sepsis involved addressing the issue of missing values. To ensure the integrity and continuity of our dataset, we adopted the “last-one carry forward” (LOCF) method for filling missing values (*Methods for handling missing data*, 2016). This method, also known as forward fill, involves filling missing data points in a time series with the last available non-null value for each variable. This approach was suitable for our datasets since the measurements were taken at regular intervals, and the last observation was a reasonable approximation for the missing value. By carrying forward the last known value, we maintained the temporal consistency of each patient’s clinical data, ensuring that our predictive models had a complete dataset to learn from.

### Feature Extraction

To enhance the predictive capability of the models for early sepsis detection, we devised a lookback window approach that was implemented for each patient. This methodology allowed the extraction of temporal statistics, providing a detailed snapshot of each patient’s physiological state over time. For each variable within this window, key statistical measures were calculated, including the maximum, mean, minimum, median, standard deviation, and the difference between consecutive measurements. We chose these statistics to encapsulate the variability and trends in the data, offering insights into the patient’s condition that are not apparent from isolated data points. In addition to these statistical features, we also generated lag features for vital signs. These lag features represented the values of vital signs at previous time points, enabling the model to incorporate information about the temporal sequence of physiological changes. This temporal sequencing was particularly relevant for sepsis prediction, where the trajectory of vital sign changes is indicative of the onset and progression of the condition. We further enhanced the dataset by including the Shock Index (SI), a vital measure of hemodynamic instability. The SI is a simple, non-invasive marker calculated as the ratio of heart rate to systolic blood pressure.

### Predictions Methodology

To facilitate a comprehensive evaluation of the predictive model, we employed an observational period that served as the foundation for our prediction strategy, delineating the historical data intervals, hereafter referred to as lookback window. These lookback intervals are crucial as they provide the temporal context from which our predictive model draws insights. For the analysis, we segmented the collected data into distinct subsets. Each subset was then processed through a specified lookback time frame. This time frame was meticulously chosen to represent the historical data depth used as input for our predictive model. To assess the predictive performance and temporal sensitivity of the model, we established a prediction horizon of six hours. This horizon represents the future time window for which the model attempts to make accurate sepsis predictions (**Figure 2**).

**Figure 2.**
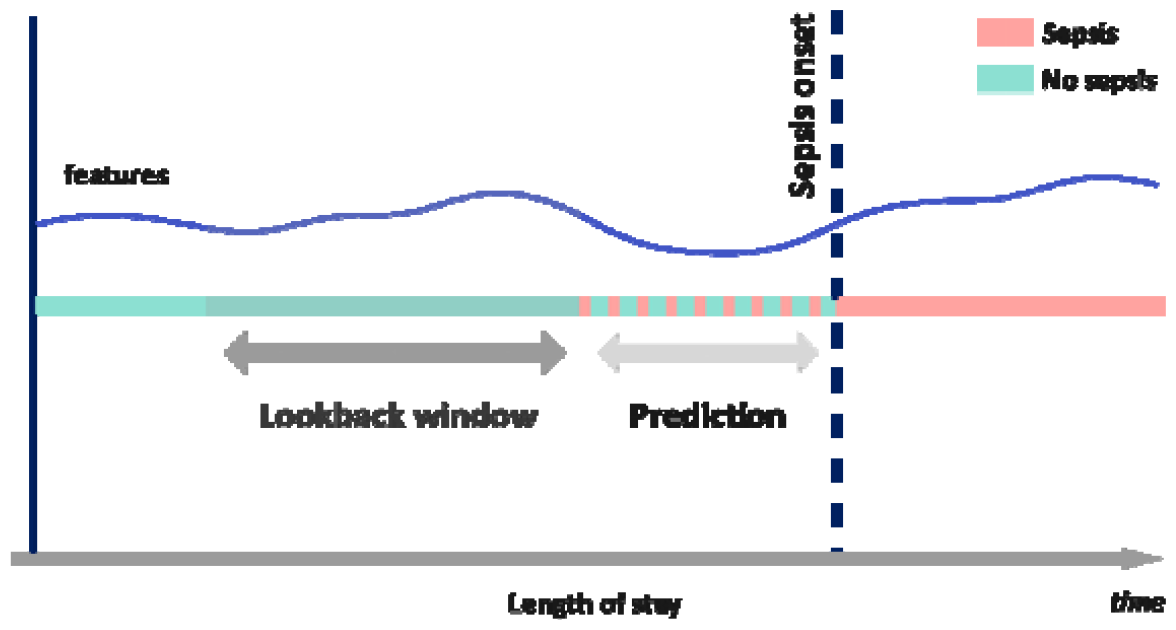
Prediction Methodology. The ‘Lookback window’ indicates the period used to gather features for the model, while the ‘Prediction’ segment shows the six-hour horizon within which the model predicts the onset of sepsis, differentiated by the red (Sepsis) and cyan (No sepsis) timelines across the patient’s length of stay.”

### Machine Learning Algorithm Development

For the prediction of sepsis onset, we utilized the XGBoost (eXtreme Gradient Boosting) algorithm. We chose XGBoost for its robustness in handling imbalanced datasets, its capability to manage missing data, and its proficiency in capturing nonlinear relationships between feature and the target variable. Prior to model training, we applied the Standard Scaler to normalize the dataset, ensuring all numerical features were standardized to have a mean of zero and a standard deviation of one. The model was trained using an enriched feature set derived, including statistical summaries and lag features of vital signs, to predict the onset of sepsis. Given the dataset’s highly unbalanced nature, we used the XGBoost *scale_pos_weight* parameter. Thi parameter is specifically designed to tackle the issue of class imbalance by adjusting the weight of the minority class, in this case, the sepsis cases, during the training of the model. We set the *scale_pos_weight* to a value that inversely reflects the proportion of the minority class. The algorithm thus compensated for the minority class underrepresentation, ensuring that the model pays more attention to correctly predicting sepsis cases.

### Hyperparameter tuning

Hyperparameters were carefully tuned to optimize model performance, balancing the trade-off between bias and variance to prevent overfitting while ensuring high predictive accuracy. For this reason, we employed Optuna, an automatic hyperparameter optimization software framework. Optuna facilitates the selection of the best set of hyperparameters by efficiently exploring the hyperparameter space using a Bayesian optimization technique. The optimization process with Optuna involved defining a search space for the hyperparameters of interest, such as learning rate, number of trees (n_estimators), depth of trees, and regularization terms. Optuna then iteratively tested different combinations of these hyperparameters in the defined search space, assessing model performance based on a predefined objective function. We aimed at maximizing the area under the receiver operating characteristic curve (AUC-ROC) to ensure both high sensitivity and specificity in sepsis prediction. Each trial in the optimization process involved training the XGBoost model with a unique set of hyperparameters and evaluating its performance using cross-validation on the training dataset.

### Performance Validation

To quantify the prediction performance of our model we computed the accuracy, precision, the F1 score, specificity, and the Area Under the Receiver Operating Characteristic (AUC-ROC) we defined the confusion matrix for the prediction as follows:

- **True Positives** (TP): Instances where the model correctly predicted the presence of sepsis.
- **False Positives** (FP): Instances where patients without sepsis were incorrectly identified by the algorithm to be at risk of developing sepsis.
- **True Negatives** (TN): Instances where the model accurately identified patients without sepsis.
- **False Negatives** (FN): Instances where the model failed to predict sepsis in patients who actually developed it.

## RESULTS

### Patient Characteristics

We analyzed data from the Physionet Cardiology Challenge 2019, focusing on adult ICU patients labeled as septic and non-septic. Patients below the age of 18 were excluded. The dataset consisted of a total of 1,259,376 observations, out of which 22,808 were labeled as septic according to Sepsis-3 criteria, revealing the challenge posed by the imbalanced nature of the clinical data. The septic and non-septic groups were nearly identical in mean age. The gender distribution in both cohorts showed a higher proportion of males, with a male-to-female ratio of approximately 1.45 in the septic group and 1.28 in the non-septic group, suggesting a slightly higher risk or a higher detection rate of sepsis among males in this ICU population. Furthermore, the length of ICU stay (in hours) demonstrates a marked difference between the two groups, with septic patients having a mean stay of nearly 65 hours, significantly longer than the 38 hours for non-septic patients. This difference is also reflected in the median values, indicating that on average, septic patients tend to stay longer in the ICU, which is an expected outcome given the complexity of sepsis management. A detailed breakdown of the demographics and clinical characteristics of the patient cohorts (both septic and non-septic groups) is presented in **Table 1**.

**Table 1.** Basic characteristics of the patient population.

| Grouped by Status (sepsis/non sepsis) |  |  |
| --- | --- | --- |
| Patient characteristic | Sepsis | Non sepsis |
| Number of observations | 22,808 | 1,259,376 |
| **Age (years)** |  |  |
| Mean | 61.84 | 61.85 |
| Median | 64 | 63.37 |
| **Gender** |  |  |
| Male | 1,415 | 16,917 |
| Female | 975 | 13,220 |
| **Length of ICU stay (hours)** |  |  |
| Mean | 64.92 | 37.98 |
| Median | 44 | 40 |

### Data preprocessing results

In our analysis, we identified that approximately 8% of the patient cohort was classified as septic, a significant finding given that explicit timestamps indicating sepsis accounted for only around 2% of the dataset. To address this imbalance and improve model performance, we leveraged the XGBoost algorithm’s *scale_pos_weight* parameter. This parameter was critical for enhancing the model’s sensitivity to the less represented septic cases, calculated based on the ratio of non-septic to septic patients, thereby ensuring a balanced consideration during training.

The dataset exhibited a notable amount of missing data, as visualized in the heatmap of **Figure 3**. The heatmap reveals dense blue areas, indicating higher data completeness, contrasted starkly with white gaps that signify the absence of recorded values across various features. The extreme rates of missing values could potentially bias our model’s predictions. Thus, features such as *Troponin, Fibrinogen*, or *Bilirubin_total* were excluded from our analysis due to high missing rate (over 95%). This step of feature elimination, along with the application of the last-one carry forward method for handling missing values in remaining variables, was instrumental in maintaining the integrity and robustness of the dataset for predictive modeling.

**Figure 3.**
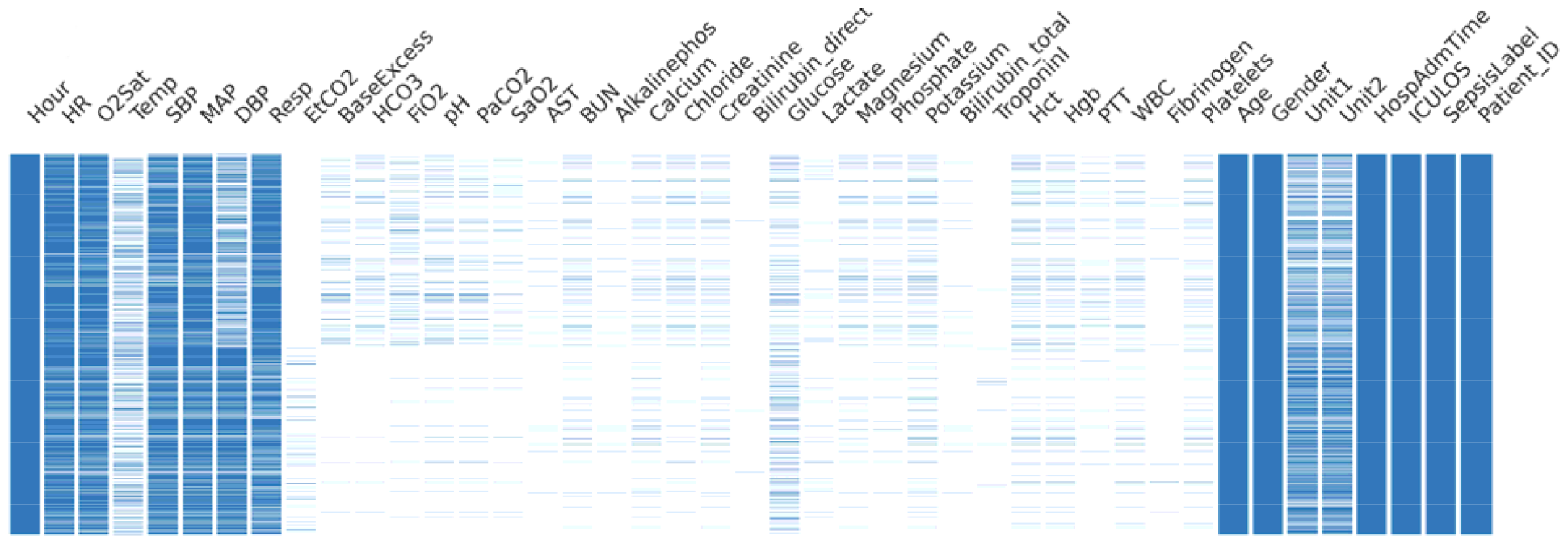
Heatmap of missing data across the dataset. The dense blue areas suggest a higher completeness of data, whereas the white gaps highlight the absence of recorded values for each feature.

### Prediction Results

Our XGBoost-based predictive model was trained on 75% of the data and optimized using Optuna under 100 trials, with the remaining 25% reserved for testing. The model’s performance was rigorously validated using a 10-fold cross-validation method to ensure generalizability across the dataset. The model demonstrated a high AUROC of 0.98, indicating strong ability to identify sepsis despite the challenges presented by the dataset. **Figure 4** illustrates the receiver operating characteristics curve of the algorithm for hold-out test data predictions. The accuracy rate was 97% in predicting sepsis up to 6 hours.

**Figure 4.**
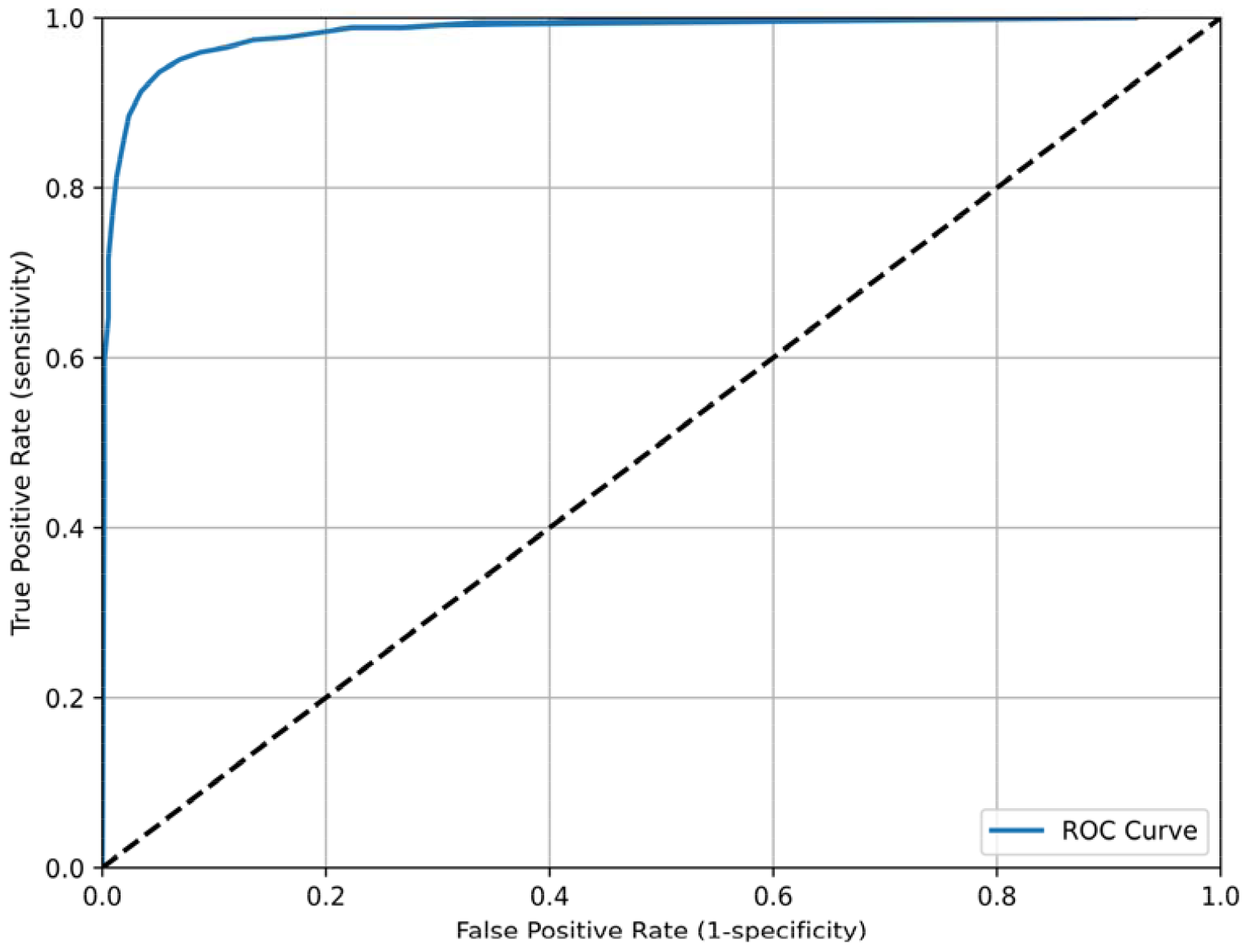
Receiver operating characteristics curve of the algorithm for hold-out test data predictions up to six hours before sepsis onset.

The sensitivity of the model, standing at 87%, underscored a high true positive rate. The model’s specificity was recorded at 98%, illustrating a high true negative rate. A key factor in refining the model’s performance was the utilization of the XGBoost’s *scale_pos_weight* parameter. Thi adjustment proved crucial for calibrating the model, especially given the complexity of the dataset due to the class imbalance. **Table 2** provides a detailed summary of the prediction results, including essential metrics such as accuracy, sensitivity, and specificity. **Figure 5** shows the confusion matrix, where the model predicted sepsis in 98.53% of the cases (Specificity, TP) with 1.47% missed cases (FN). The model correctly predicted that the patients did not develop sepsis in 86.53% (Sensitivity, TN) with 13.47% of the cases predicted falsely as sepsis (FP).

**Table 2.**
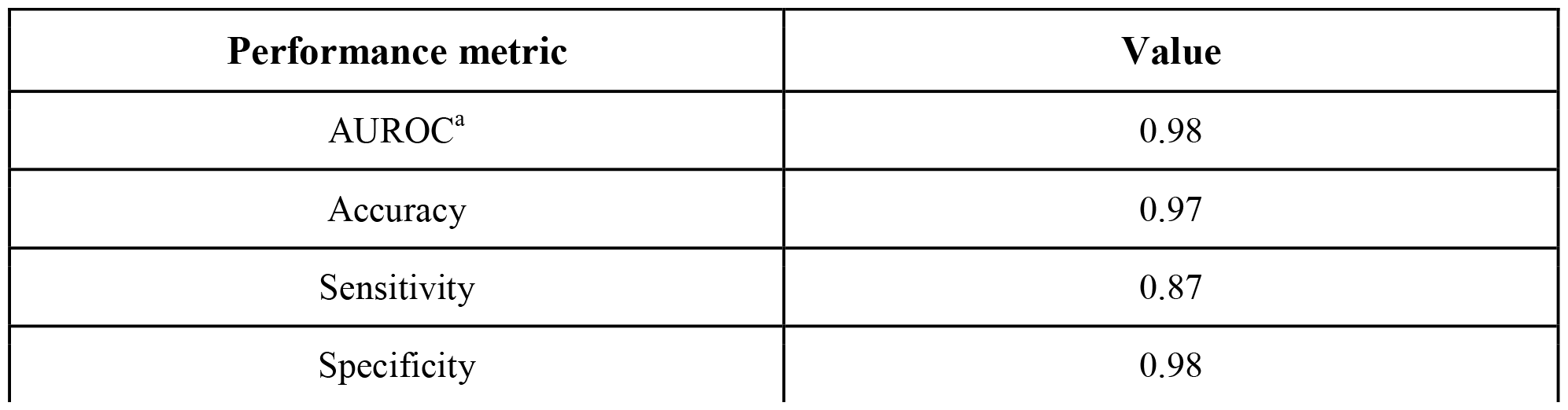
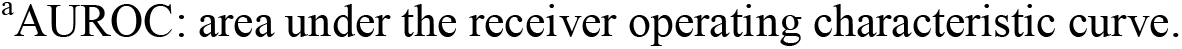
XGBoost performance on test data.

| Performance metric | Value |
| --- | --- |
| AUROC<sup>a</sup> | 0.98 |
| Accuracy | 0.97 |
| Sensitivity | 0.87 |
| Specificity | 0.98 |

<sup>a</sup>AUROC: area under the receiver operating characteristic curve.

**Figure 5.**
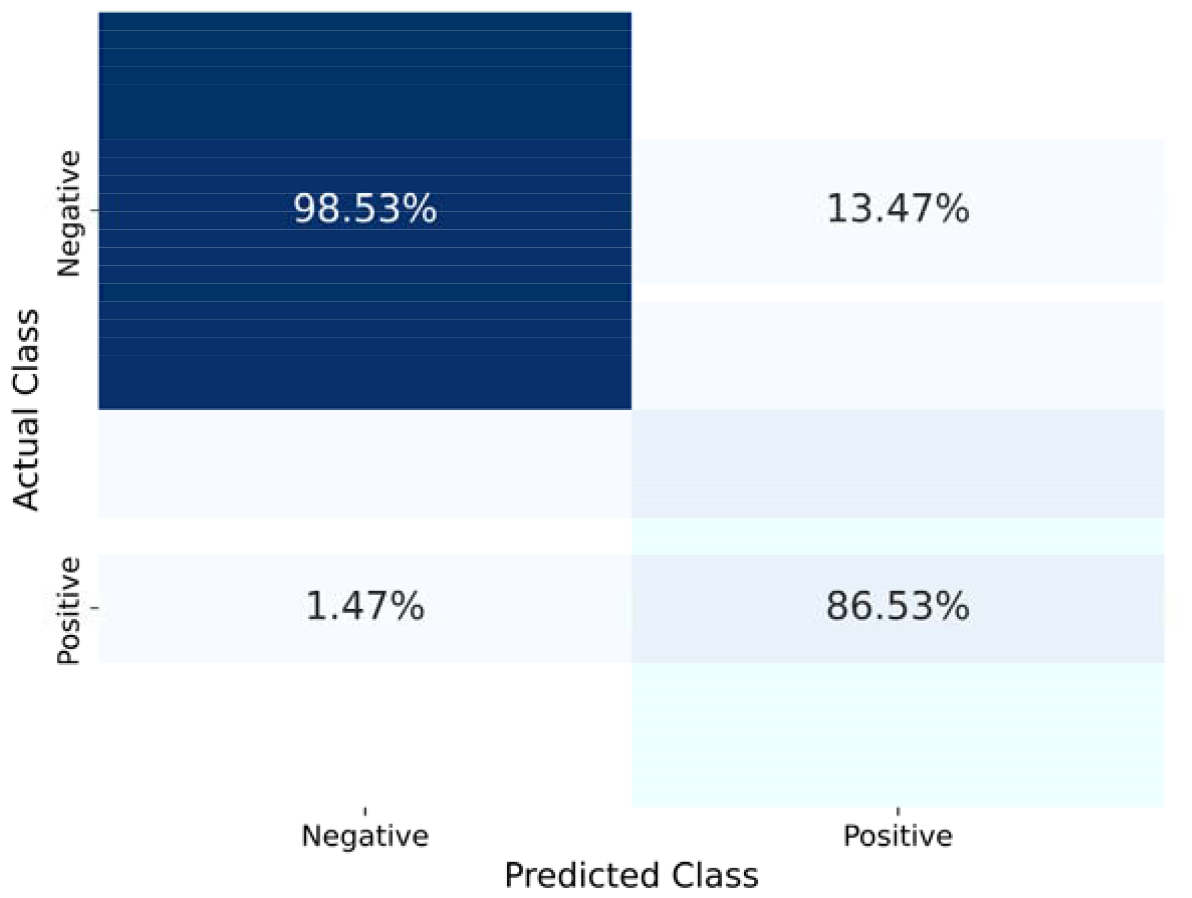
Confusion matrix of the XGBoost model.

### Comparison to previous work

We compared the performance of our AI model with other clinical screening tools (i.e. MEWS, SOFA, and SIRS) as well as with commercially available AI products to demonstrate the potential application of VIOSync’s SPI in the clinical practice (**Figure 6**). VIOSync’s SPI achieved an AUROC = 0.98 that was higher than SOFA (AUROC = 0.83), NEWS (AUROC = 0.81) and SIRS (AUROC = 0.82). Interestingly qSOFA has worse performance than SOFA with an AUROC = 0.8. With respect to other commercial softwares, the platform offered by BIOcogniv achieves similar performance to our VIOSync’s SPI performance with an AUROC = 0.94. The commercial model with the worst performance was the NAVOY algorithm by AlgoDx that achieved an AUROC = 0.8. The AUROC scores for the commercially available models have been extracted from either peer-reviewed publications of the companies or their commercial websites. To our knowledge we used the latest sources that reported these values. As such there may exist potential deviations from their performance that could be currently validated in clinical trials or larger cohorts of patients.

**Figure 6.**
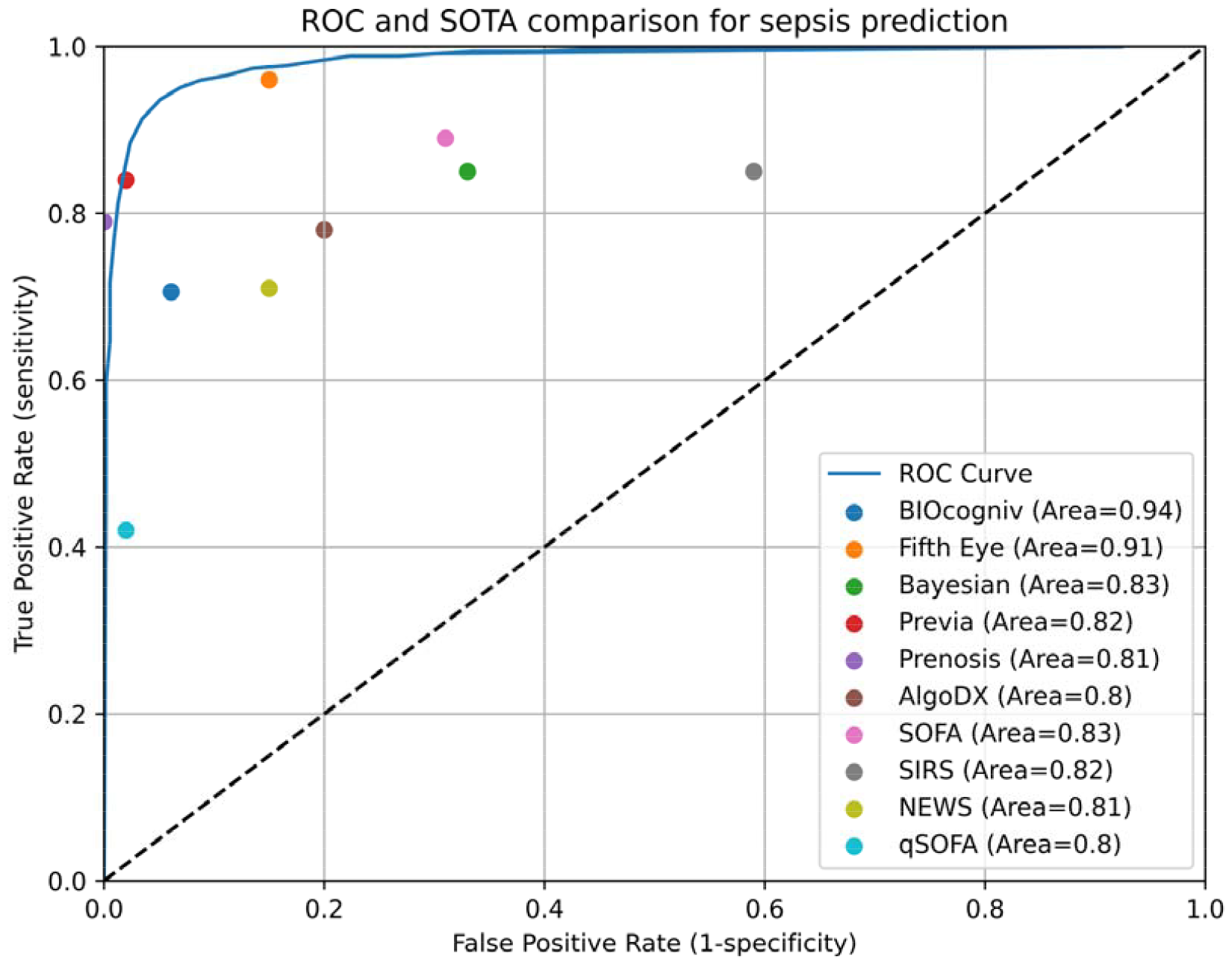
ROC curves of the VIOSync SPI model compared to SOTA model and scores for sepsis prediction onset.

## DISCUSSION

### Principal Results

Early identification and treatment of sepsis is a highly complex and multifaceted challenge and requires highly skilled and well-trained human experts (Helms and Perner, 2020; Komorowski, 2020). This study used a supervised machine learning method to build a predictive model of sepsis events predicted by XGBoost. The dataset’s inherent imbalance, with a significant majority of non-septic instances, presented a substantial challenge toward this goal. Despite that, the sensitivity, specificity and AUC of this proposed method was 87%, 98%, and 0.98, respectively, demonstrating excellent predictive performance. The SPI algorithm more accurately predicted the onset of sepsis developed during hospitalization than the frequently used rules-based patient decompensation screening tools MEWS, SOFA, and SIRS. While used for sepsis screening in many clinical settings, these tools are not designed to exploit information from trends in patient data, and demonstrate suboptimal efficiency (Vincent *et al*., 1996; Subbe *et al*., 2006; Usman, Usman and Ward, 2019). In comparison to other predictive models, the SPI algorithm’s performance metrics not only surpass those of MEWS, SOFA, and SIRS but also exhibit competitive advantage to other leading machine learning models in the field. This includes models based on deep learning, neural networks, and other ensemble methods that have been proposed for sepsis prediction. **Figure 6** provides a visual representation of this comparative analysis, plotting the AUROC of the SPI algorithm against both the traditional risk scores like qSOFA and SIRS and other SOTA models. The graph clearly demonstrates the superior performance of the SPI algorithm, with its AUC higher than that of the conventional tools and comparable or superior to the AUCs of other advanced predictive models.

### Limitations

This study has some limitations. First, the algorithm was developed using retrospective data and has not yet been evaluated prospectively. Second, it would have been valuable to test the performance of the algorithm with an additional external validation cohort, for example, data from the MIMIC III (Goldberger *et al*., 2000; Johnson *et al*., 2016) or IV (Goldberger *et al*., 2000; Johnson *et al*., 2023), or the eICU Collaborative Research Database (Goldberger *et al*., 2000; Pollard *et al*., 2018). It should, however, be noted that external validation was performed in this study on hold-out test data. Because we do not perform any subgroup analyses in the present study, we also cannot verify the generalizability of these results to specific patient subpopulations. Future work investigating performance on subpopulations defined by medical or demographic characteristics is therefore warranted. Moreover, this study does not provide information on the clinical or economic impact of the integration of the developed algorithm in clinical practice. Finally, because our study is a retrospective analysis of encounters which do not involve the intervention of predictions from the SPI algorithm, we must await real-time, prospective evaluation of the algorithm before making claims of impact on clinical practice and patient outcomes.

### Future Work

The accuracy, sensitivity, and specificity of the SPI algorithm developed in this study are to potentially be validated in a prospective clinical trial (ClinicalTrials.gov; NCT06238180). That study also intends to further explore the developed algorithm’s integration into clinical workflow and effect on relevant clinical outcomes beyond the ICU (i.e., surgical ward). Finally, with access to data from different institutions, the algorithm can be retrained and continuously improved or adjusted to work well in different settings (regions, hospitals, populations).

## Conclusions

Sepsis remains a leading cause of mortality and morbidity in ICUs worldwide. Early detection is key to effective management and patient outcome, as there is no specific sepsis treatment available. We have developed a high-performance machine learning sepsis prediction algorithm that outperforms existing early warning scoring systems. The algorithm is based on variables routinely collected and readily available in electronic health records in ICUs of all categories and may provide an opportunity for enhanced patient monitoring, earlier detection of sepsis, and improved patient outcomes. Should the results of this study be confirmed by future prospective randomized clinical trials, our algorithm has the potential to emerge as a groundbreaking tool for use in hospitals, establishing a new benchmark for early detection of sepsis. A tool like VIOSync SPI, which identifies patients at risk of sepsis early, would offer caregivers a greater opportunity to intervene before clinical deterioration and the onset of sepsis.

## Data Availability

All data produced in the present work are contained in the manuscript

## Author contributions

Conceptualization: SGL, VSD, IAG

Methodology: SGL

Results Interpretation: VSD, IAG, AD

Project administration: VSD, IAG

Planning and Supervision: VSD, IAG

Writing – original draft: SGL, VSD, IAG

Writing – review & editing: SGL, VSD, IAG, AD

## Acknowledgments

We thank Agathi Kari for her initial help on setting-up the preprocessing pipeline of the current study.

